# Prevalence of SARS-CoV-2 infection among asymptomatic healthcare workers in greater Houston: a cross-sectional analysis of surveillance data from a large healthcare system

**DOI:** 10.1101/2020.05.21.20107581

**Authors:** Farhaan S. Vahidy, H. Dirk Sostman, David W. Bernard, Marc L. Boom, Ashley L. Drews, Paul Christensen, Jeremy Finkelstein, Bita A. Kash, Robert A. Phillips, Roberta L. Schwartz

## Abstract

**Objective:** To determine the prevalence of SARS-CoV-2 infection among asymptomatic COVID-19 facing and non-COVID-19 facing Healthcare Workers (HCWs), with varying job categories across different hospitals.

**Design:** Cross-sectional analysis of a healthcare system surveillance program that included asymptomatic clinical (COVID-19 facing and non-COVID-19 facing), and non-clinical HCWs. A convenience sample of asymptomatic community residents (CR) was also tested. Proportions and 95% confidence Intervals (CI) of SARS-CoV-2 positive HCWs are reported. Proportional trend across HCW categories was tested using Chi Square trend test. Logistic regression model-based likelihood estimates of SARS-CoV-2 prevalence among HCWs with varying job functions and across different hospitals are reported as adjusted odds ratios (aOR) and CI.

**Setting:** Healthcare system comprising one tertiary care academic medical center and six large community hospitals across Greater Houston and a community sample.

**Participants:** 2,872 self-reported asymptomatic adult (> 18 years) HCWs and CRs.

**Exposure:** Clinical HCWs in COVID-19 and non-COVID-19 units, non-Clinical HCWs, and CRs. Job categories of Nursing, Providers, Allied Health, Support, and Administration / Research. Seven hospitals in the healthcare system.

**Main Outcomes:** Positive reverse transcriptase polymerized chain reaction (RT-PCR) test for SARS-CoV-2

**Results:** Among 2,872 asymptomatic HCWs and CRs, 3.9% (CI: 3.2 – 4.7) tested positive for SARS-CoV-2. Mean (SD) age was 40.9 (11.7) years and 73% were females. Among COVID-19 facing HCWs 5.4% (CI: 4.5 – 6.5) were positive, whereas 0.6% (CI: 0.2 – 1.7%) of non COVID-19 facing HCWs and none of the non-clinical HCWs or CRs were positive (*P_trend_* < 0.001). Among COVID-19 facing HCWs, SARS-CoV-2 positivity was similar for all job categories (p = 0.74). However, significant differences in positivity were observed across hospitals.

**Conclusions and Relevance:** Asymptomatic HCWs with COVID-19 patient exposure had a higher rate of SARS-CoV-2 positive testing than those not routinely exposed to COVID-19 patients and those not engaged in patient care. Among HCWs with routine COVID-19 exposure, all job types had relatively similar infection rates. These data can inform hospital surveillance and infection control practices for patient-facing job classifications and suggest that general environmental exposure within hospitals is not a significant source of asymptomatic SARS-CoV-2 infection.

**What is already known on this topic:**

- A sizeable proportion of individuals who contract the novel Severe Acute Respiratory Syndrome Coronavirus 2 (SARS-CoV-2) can remain largely asymptomatic.
- Though such individuals may not develop symptoms, they continue to shed enough viral particles to trigger positive reverse transcriptase polymerized chain reaction (RT PCR) test for SARS-CoV-2
- Prior reports on proportion of asymptomatic SARS-CoV-2 individuals are highly variable with positivity ranging across < 1% to 36%
- Asymptomatic SARS-CoV-2 infection among healthcare workers is specifically critical to understand

**What this study adds:**

- This study demonstrates that overall rate of SARS-CoV-2 infection among asymptomatic healthcare workers in a large healthcare system of a metropolitan city in the United States was 3.9%
- The rate of SARS-CoV-2 infection among healthcare workers who provided direct care to COVID-19 patients was 5.4% whereas it was 0.6% among those healthcare workers who did not provide direct care to COVID-19 patients
- There was no difference in SARS-CoV-2 positivity rate for different job categories of healthcare workers who provided direct care to COVID-19 patients

## INTRODUCTION

Transmission of the SARS-CoV-2 virus by asymptomatic individuals is a public health concern. Estimates of asymptomatic spread in the population are highly variable (6% - 79%).^1-3^ Healthcare workers (HCWs) are at a higher risk for infection and can become inadvertent vehicles of transmission.^4,5^ Therefore, characterizing asymptomatic SARS-CoV-2 infection within healthcare organizations is critical. Houston Methodist (HM) initiated a coronavirus disease-2019 (COVID-19) surveillance program among asymptomatic HCWs and expanded it to asymptomatic community resident (CR) volunteers. We report prevalence of SARS-CoV-2 among the first group tested.

## METHODS

### Study Design and Population

Our system comprises an academic medical center (AMC), six acute care community hospitals and a continuing care hospital throughout greater Houston that all provided care to COVID-19 patients.

The HCWs included 3 sub-groups: 1) clinical workers in patient care areas with COVID-19 patients, 2) clinical workers in segregated units without COVID-19 patients, and 3) non-clinical workers with no patient contact. Since within clinical units certain job categories may have greater intensity or duration of patient exposure; in a secondary analysis, we classified study participants who worked in COVID-19 patient care units into five job categories. These were: 1) *Nursing* (bed side registered nurses [RNs], nurse aides, bedside technicians, and emergency medical technicians), 2) *Providers* (physicians, residents, nurse practitioners, physician assistants), 3) *Allied Healthcare Workers* (therapists, non-bedside technicians, pharmacists, social workers), 4) *Support staff* (housekeeping, security), and 5) Administrative / Research (managers, coordinators, administrative assistants, research staff).

### Data Collection and Ethical Approvals

We collected nasopharyngeal swabs, age, and sex information from self-reported asymptomatic HCW volunteers and consented CRs, under a surveillance initiative for HCWs and an IRB approved protocol for CRs. Testing for SARS-CoV-2 was done by one of the three cross-validated reverse transcriptase polymerase chain reaction (RT-PCR) assays. Participants were informed of their test results.

### Statistical Analysis

We report proportions and 95% confidence intervals (CI) for SARS-CoV-2 infection among study sub-groups. Means (SD) for age and proportional distribution of sex for these categories are also provided. Proportional test for trend^6^ was used to explore the association between RT-PCR positivity and various HCW sub-groups. We fitted logistic regression models,^7^ and report adjusted odds ratios (aOR) and CI for likelihood of positive SARS-CoV-2 infection across seven hospitals and five job categories within the COVID-19 facing HCWs. For our primary comparison of COVID-19 facing and non-COVID-19 facing HCWs, we anticipated a 3% absolute difference (5% among COVID-19 facing and 2% among non-COVID-19 facing HCWs) in positive SARS-CoV-2 proportion. First phase testing was planned for at least 2,500 patients that provided > 90% power to test the anticipated difference with type I error rate of 5%.

### Patient and Public Involvement

Neither patients nor the public were involved in the conception or conduct of the study.

## RESULTS

A total of 2,872 individuals (2,787 HCWs and 85 CRs) were included. Overall, the mean (SD) age was 40.9 (11.7) years and 73% were females. Table 1 shows age and sex distribution for CRs and various HCW categories.

**Table 1:**
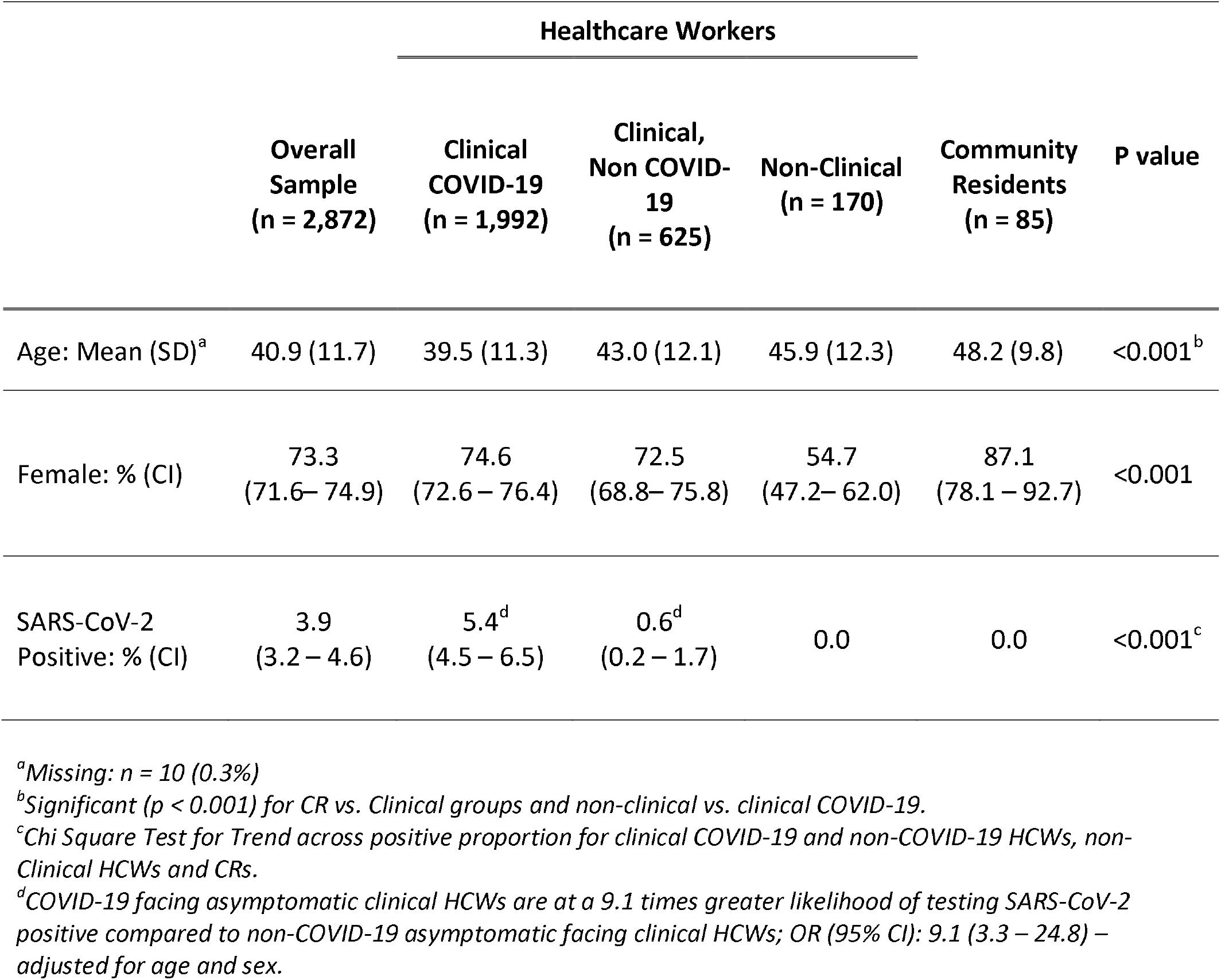
Age, Sex and SARS-CoV-2 Positive Proportions Among Three Categories of Asymptomatic Healthcare Workers and Community Residents

The overall SARS-CoV-2 positive proportion (CI) in our study population was 3.9% (3.2 – 4.7). Among the clinical HCWs, 5.4% (4.5 – 6.5) from COVID-19 units tested positive, whereas 0.6% (0.2 – 1.7) from non-COVID units and none of the non-clinical HCWs or CRs tested positive (*P_trend_* < 0.001). After adjusting for age and sex, this represents a 9 times higher likelihood of SARS-CoV-2 positivity among asymptomatic COVID-19 facing HCWs, compared to non-COVID-19 facing HCWs, aOR (95% CI): 9.1 (3.3 – 24.8) (Table 1).

Among asymptomatic HCWs in COVID-19 patient care units (n = 1,992), the proportion of SARS-CoV-2 positivity ranged between 3.6% (CI: 1.3 – 9.1) for support staff to 6.5% (CI: 3.9 – 10.7) for allied health and 6.5% (CI: 3.6 – 11.3) for administrative / research staff. However, the proportions of SARS-CoV-2 positivity were not significantly different across the five job categories of COVID-19 facing asymptomatic HCWs (P*_trend_* = 067). The statistical non-significance across various job categories continued to be observed after adjusting for potential differences in age and sex distribution among COVID-19 facing HCWs. Table 2 provides summary of age, sex, and SARS-CoV-2 positive distribution across various job categories for COVID-19 facing HCWs.

**Table 2:** Age, Sex, and SARS-CoV-2 Positive Proportions Among COVID-19 Facing Clinical Healthcare Workers across different job categories

|  | **Nursing**<br>**(n = 1,279)** | **Provider**<br>**(n = 210)** | **Allied Health**<br>**(n = 214)** | **Support**<br>**(n = 112)** | **Admin /**<br>**Research**<br>**(n = 170)** | **P value** |
| --- | --- | --- | --- | --- | --- | --- |
| Age:<br>Mean (SD) | 38.1 (10.8) | 41.5 (9.8) | 39.1 (11.4) | 45.4 (13.6) | 44.6 (11.4) | < 0.001<sup>a</sup> |
| Female:<br>% (CI) | 79.0<br>(76.6 – 81.1) | 52.4<br>(45.6 – 59.1) | 65.4<br>(58.8 – 71.5) | 76.8<br>(68.1 – 83.7) | 78.2<br>(71.4 – 83.8) | < 0.001 |
| SARS-CoV-2 Positive:<br>% (CI) | 5.2<br>(4.1 – 6.6) | 5.7<sup>c</sup><br>(3.3 – 9.8) | 6.5<sup>c</sup><br>(3.9 – 10.7) | 3.6<sup>c</sup><br>(1.3 – 9.1) | 6.5<sup>c</sup><br>(3.6 – 11.3) | 0.67<sup>b</sup> |

<sup>a</sup>Statistically significant ( $p < 0.001$ ) for Nursing and Allied Health Categories vs. Other categories
<sup>b</sup>Chi Square Test for Trend across job categories
<sup>c</sup>Adjusted OR (95% CI) for Provider: 1.1 (0.6 – 2.0), Allied Health: 1.2 (0.7 – 2.3), Support: 0.6 (0.2 – 1.7), and Admin / Research: 1.2 (0.6 – 2.3), with Nursing as reference category, adjusted for age and sex.

We further compared proportions SARS-CoV-2 positive asymptomatic COVID-19 facing HCWs across various hospitals in our healthcare system; and observed considerable variation in the range of SARS-CoV-2 positivity from no infection to 12.5% positive proportion. After adjusting for age, sex and job category, two hospitals demonstrated significantly higher likelihood of SARS-CoV-2 positivity among asymptomatic COVID-19 facing HCWs compared to the Academic Medical Center, aOR (95% CI): 2.78 (1.76 – 4.39) and 2.49 (1.23 – 5.02), whereas infection rate was significantly lower in another facility compared to the central tertiary care hospital, aOR (95% CI): 0.34 (0.12 – 0.95). The output of the fully adjusted logistic regression model is provided in Table 3.

**Table 3:**
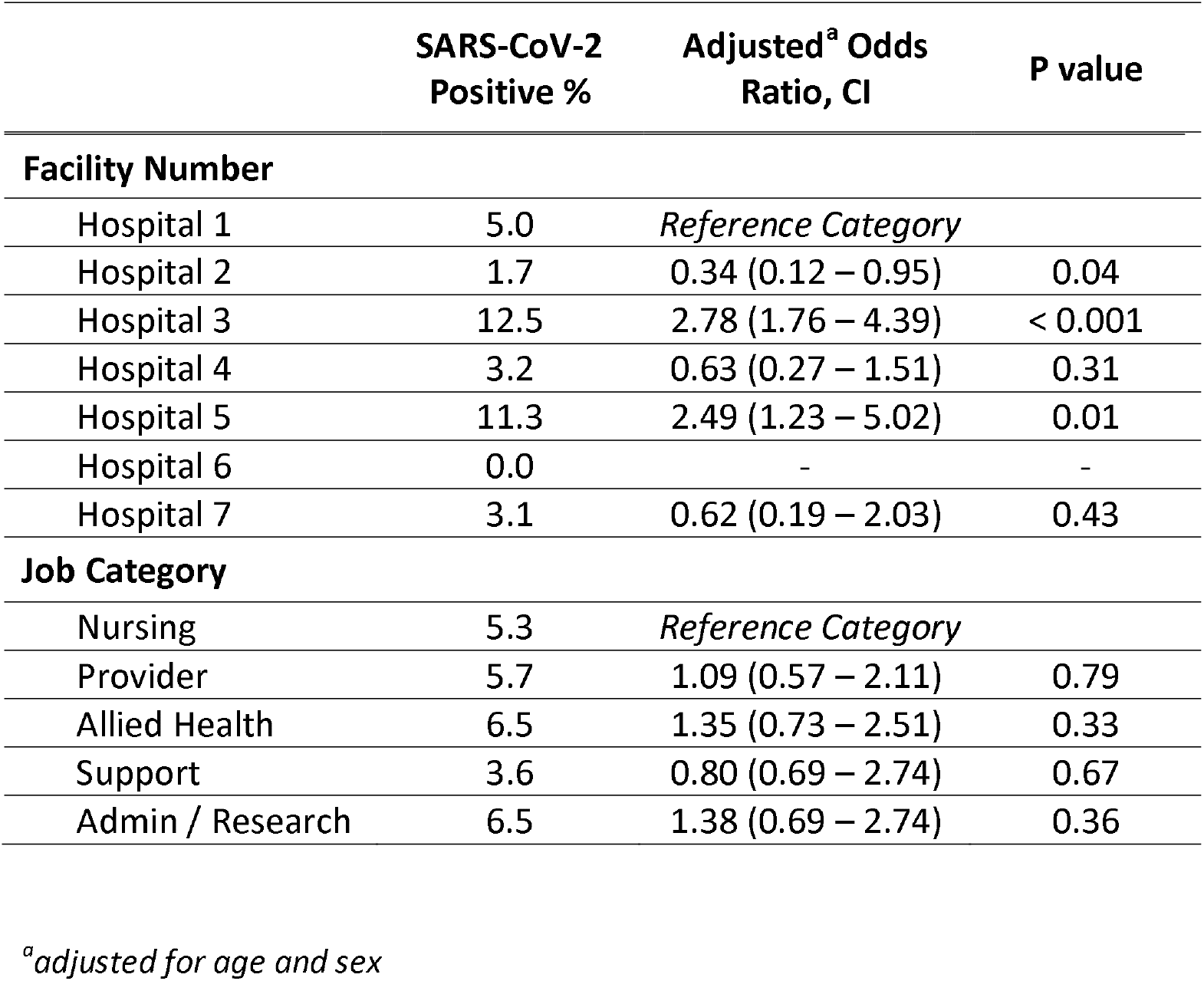
Multivariable Model Based Likelihood Estimates of SARS-CoV-2 Positive Results Across Different Hospitals in Various Job Categories for Asymptomatic COVID-19 Facing Healthcare Workers

## DISCUSSION

As the COVID-19 pandemic continues to unfold and strategies for ‘re-opening’ are contemplated; understanding asymptomatic transmission of SARS-CoV-2 among HCWs is vitally important.^8-11^ Though asymptomatic SARS-CoV-2 prevalence among general population has been reported^10-12^ to our knowledge, this is the first report describing asymptomatic SARS-CoV-2 infection among HCWs in a large U.S. metropolitan healthcare system.

To evaluate which HCWs and hospital units are at a higher risk for asymptomatic virus transmission, we compared SARS-CoV-2 positive proportion among COVID-19 facing and non-COVID-19 facing clinical employees. The statistically significant difference of 4.8% between these two groups and a low overall prevalence among non-COVID facing HCWs (0.6%); coupled with the absence of infection in the non-clinical HCWs and the CRs is consistent with higher transmission from patients or co-workers among HCWs in COVID-19 units, rather than community transmission or acquisition of infection from the general hospital environment.^11,12^ Across our healthcare system, non-clinical HCWs worked in buildings which did not involve direct patient care and had separate entrances and HVAC systems. Furthermore, population density in these buildings was low since most assigned there were working remotely, as per institutional policies. Similarly, the sample of CRs was drawn from individuals with no exposure to the hospital environment.

We performed a secondary analysis of prevalence of SARS-CoV-2 infection across various job categories among COVID-19 facing HCWs. This comparison did not yield a statistically significant difference between presumably high and low exposure job types; supporting the need for uniform infection control practices within patient care units. We speculate that simple droplet models of environmental transmission may not suffice to model exposure risks for HCWs in COVID-19 patient care units. However, we did find differences in infection rates across different hospitals within our healthcare system. Despite having uniform infection control and hygiene policies across the healthcare system, these differences may indicate variability in implementation and adherence to COVID-19 specific infection control practices. As demonstrated in our main analyses, proportion of SARS-CoV-2 positivity among non-COVID-19 facing and non-clinical HCWs was consistently low to zero. Systematic hospital factors that may lead to higher rates of SARS-CoV-2 infection among asymptomatic HCWs warrant further evaluation.

Prior reports indicate higher prevalence of asymptomatic SARS-CoV-2 infection among the general population than we found in a small sample of non-clinical HCWs and CRs.^8,11^ This could reflect a lower population prevalence in Houston or simply be a sampling variation. Although we are unaware of reports of asymptomatic HCW infection rates, an early study from Wuhan^13^ reported that 3.5% of COVID-19 patients were healthcare personnel. Furthermore, reports from Singapore, Iceland, and Italy demonstrated pre-symptomatic transmission rates of 6.4% - 43%.^3,14,15^ With a potentially high level of both asymptomatic and symptomatic patients within a hospital facility, the addition of approximately 5% asymptomatic infections in HCWs would have implications for patient and employee protection, particularly for large and complex healthcare organizations. It is noteworthy that, for COVID-19 facing clinical workers, neither job category nor work location seem to guarantee safety from infection, as demonstrated recently in another study.^16^ With effective infection control measures and effective policies of managing positive individuals,^14-16^ it is thought that return to normal clinical operations can be conducted safely.

Though overall powered to demonstrate a statistically significant and clinically meaningful 3% difference in SARS-CoV-2 positivity between COVID-19 facing and non-COVID-19 facing HCWs, our findings are limited by convenience sampling from a single healthcare system and a small homogenous sample of CRs. Due to the nature of our study design, potential ‘volunteer bias’ cannot be ruled out. Furthermore, we did not control for differences in shifts and potential behavioral issues such as test avoidance for fear of lost shift work income. Additionally, in the secondary analysis, job categories were determined based on employment and do not necessarily represent a differential exposure to COVID-19 patient. The job-based comparisons may also have been underpowered.

Although, some hospitals had higher prevalence of infection in this sample, the generalizability of this findings is unclear, warranting further evaluation. However, our main findings are clear. HCWs in COVID-19 facing patient care units are at greater risk of asymptomatic infection compared to other HCWs or CRs. Accordingly, it is likely that these infections are acquired within these patient care units rather than in the community or in the common areas of the hospital such as hallways or cafeterias. Within COVID-19 patient care units, all workers have similar risks of having asymptomatic infection. Infection control practices within hospitals should account for these differences.

## CONCLUSIONS

In this study of asymptomatic HCWs, the higher rate of positive test results in those with COVID-19 patient exposure highlights the need for hospitals to implement surveillance and infection control policies, regardless of an employee’s job classification. The differences we noted between hospitals also highlights the need for consistent practices throughout an organization. It seems likely that ongoing surveillance of the healthcare workforce will be essential during the COVID-19 outbreak and is of particular importance when efforts are made to restore normal clinical operations.

## Data Availability

De-identified data may be available by contacting the corresponding author, contingent upon institutional approvals.

## Acknowledgements

All authors declare no conflict of interest. We gratefully acknowledge the superb writing skills of Dr. Kimberly Greer, as well as the staff in the Houston Methodist Clinical Laboratory who performed all testing, and the infection control staff led by Firas Zabaneh who work tirelessly to help keep us all safe. And of course, the front-line health care workers and providers have our utmost respect and admiration; without them, this work would have been not only impossible, but bereft of meaning.

